# Reliability of wastewater analysis for monitoring COVID-19 incidence revealed by a long-term follow-up study

**DOI:** 10.1101/2021.05.30.21257992

**Authors:** Rafael Sanjuán, Pilar Domingo-Calap

## Abstract

**Background:** Wastewater-based epidemiology has been used for monitoring human activities and waterborne pathogens. Although wastewaters can also be used for tracking SARS-CoV-2 at the population level, the reliability of this approach remains to be established, especially for early warning of outbreaks.

**Methods:** We collected 377 samples from different treatment plants processing wastewaters of >1 million inhabitants in Valencia, Spain, between April 2020 and March 2021. Samples were cleaned, concentrated, and subjected to RT-qPCR to determine SARS-CoV-2 concentrations. These data were compared with cumulative disease notification rates over 7 and 14 day periods.

**Results:** We amplified SARS-CoV-2 RNA in 75% of the RT-qPCRs, with an estimated detection limit of 100 viral genome copies per liter (gc/L). SARS-CoV-2 RNA concentration correlated strongly with disease notification rates over 14-day periods (Pearson *r* = 0.962, *P* < 0.001). A concentration >1000 gc/L showed >95% sensitivity and specificity as an indicator of more than 25 new cases per 100,000 inhabitants. Albeit with slightly higher uncertainty, these figures were reproduced using a 7-day period. Time series were similar for wastewaters data and declared cases, but wastewater RNA concentrations exhibited transient peaks that were not observed in declared cases and preceded major outbreaks by several weeks.

**Interpretation:** Wastewater analysis provides a reliable tool for monitoring COVID-19, particularly at low incidence values, and is not biased by asymptomatic cases. Moreover, this approach might reveal previously unrecognized features of COVID-19 transmission.

**Funding:** Conselleria d’Agricultura, Desenvolupament Rural, Emergencia Climàtica i Transició Ecològica of the Generalitat Valenciana (project OTR2020-20593SUBDI), Instituto de Salud Carlos III (FONDO-COVID19 COV20/00210), CSIC (Salud Global CSIC 202020E292), and Ministerio de Ciencia e Innovación (Ramón y Cajal contract, Call 2019).

## Introduction

The COVID-19 pandemic has enforced severe epidemiological control measures in many countries, including restrictions on population movement, lockdowns, and reduced social and economic activities. SARS-CoV-2 is an airborne virus that replicates in the respiratory tract, but also in other body compartments such as the intestinal mucosa. As a result, viral material is excreted through feces and other specimens such as urine or saliva.^1,2^ This offers the possibility of monitoring the presence of SARS-CoV-2 at the population level using wastewater analysis.^3– 14^ Previous work has carried out short-term follow-ups including treatment plants in the Frankfurt metropolitan area (Germany) from March to September 2020^15^, in San Diego (USA) from July to October 2020^16^, in Brisbane, Queensland (Australia) from February to May 2020^17^, in Paris (France) from March to April 2020^18^, in Santa Catarina (Brazil) from October 2019 to March 2020^19^, and in Barcelona (Spain) from April to July 2020.^20^

These works suggest that wastewater analysis can provide useful epidemiological information. However, whether COVID-19 prevalence can be reliably inferred from wastewaters is an open question. At present, the observed correlation between viral RNA concentration in wastewaters and declared COVID-19 cases has not been high enough to achieve this goal.^21,22^ This might reflect intrinsic limitations to the method or lack of sufficient sampling effort. Another open question is whether wastewaters can anticipate COVID-19 incidence trends and, thus, can serve as an early warning tool.^23,24^ Some studies suggest an anticipation of 2-4 days based on short-term follow-ups, with variations depending on the period under study.^25,26^ However, a caveat concerning these analyses is that COVID-19 diagnosis was underreported during the early stages of the pandemics, since systematic PCR testing of suspected cases was not efficiently implemented at that time. For instance, high levels of viral RNA were found in wastewaters collected during early March 2020 in the metropolitan area of Valencia, Spain, whereas declared cases lagged weeks behind due to poor testing.^12^ Finally, wastewaters might reveal novel aspects of COVID-19 epidemiology. Since approximately half of the infections are asymptomatic and disease severity is strongly age-dependent, large fractions of young populations are likely to escape diagnosis, and wastewater analysis could in principle help us correct this detection bias.

Here, we use wastewaters to carry out a nearly one-year follow-up study of SARS-CoV-2 incidence in a large city. To achieve this goal, we monitored treatment plants in the metropolitan area of Valencia, Spain, from April 2020 to March 2021. We found that SARS-CoV-2 RNA concentration in wastewaters was strongly correlated with the number of declares cases over 7- and 14-day periods. Consequently, wastewaters provided an accurate indicator of COVID-19 incidence. Throughout multiple epidemic waves, we found no conclusive evidence supporting the ability of SARS-CoV-2 RNA concentration in wastewaters to systematically anticipate an increase in the number of declared cases. Nevertheless, wastewater SARS-CoV-2 RNA concentrations experienced transient peaks preceding such waves, a unique feature that was not revealed by the analysis of declared cases, and that warrants future investigation.

## Results

We obtained composite wastewater samples from the wastewater treatment plant (WWTP) of Pinedo, which processes sewage from 1.2 million inhabitants in the metropolitan area of Valencia, Spain. Each composite sample consisted of a pool of multiple discrete samples taken at regular intervals over a 4 h period (7-11am). Sampling was carried out on 129 different days spanning from April 16, 2020, to March 9, 2021, a period during which three recognizable epidemic waves occurred. We did not include the first outbreak that took place from February to March 2020 since PCR testing was not sufficiently implemented at that time, leading to severe case underreporting. Each sampling day, two different WWTP collectors were analyzed. Samples were spiked with the transmissible gastroenteritis coronavirus (TGEV) as an internal control, spun to remove large debris, filtrated, concentrated by high-speed centrifugation, and used for RNA extraction. We then performed SARS-CoV-2 RT-qPCR in duplicate for each of two viral genome regions (N1 and N2), and used standard curves for estimating the concentration of SARS-CoV-2 RNA in wastewater, measured as viral genome copies per L (gc/L). We successfully amplified a PCR product in 771 out of 1024 total reactions (75.3%). Failures occurred typically when the number of declared COVID-19 cases in the region was low (Fig. 1A). Moreover, RT-qPCR yielded a PCR product for TGEV in these SARS-CoV-2-negative samples, showing that lack of SARS-CoV-2 amplification was not due to issues in sample processing or the presence of PCR inhibitors. Consequently, these negative PCRs provided information about disease incidence. Analysis of the distribution of the estimated SARS-CoV-2 RNA concentrations suggested a detection limit of approximately 100 gc/L (Fig. 1B). We therefore assumed this lower-limit value for negative PCRs.

**Figure 1.**
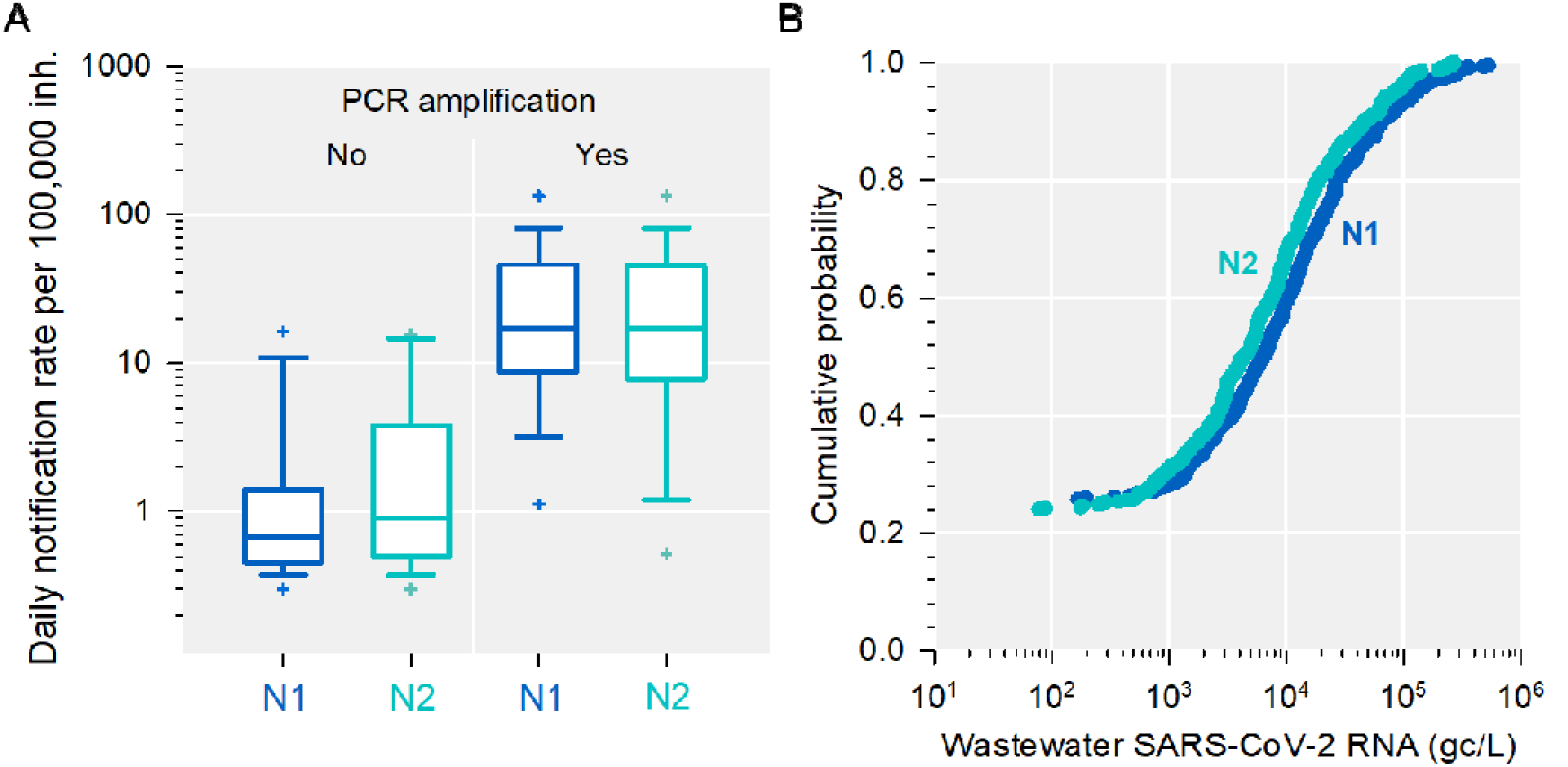
SARS-CoV-2 detection limit in wastewaters. **A**. Box plot of daily notification rates per 100,000 inhabitants for wastewater samples that yielded an RT-qPCR product, versus those that failed to amplify. Data are shown separately for viral genome regions N1 and N2. Boxes show the 25^th^, 50^th^ (median), and 75^th^ percentiles. Error bars indicate the 10^th^ and 90^th^ percentiles, and crosses show the 5^th^ and 95^th^ percentiles. **B**. Cumulative probability distribution of the estimated viral RNA concentration for genome regions N1 and N2. Based on this distribution, the detection limit of the RT-qPCR was approximately 100 gc/L.

We averaged log wastewater SARS-CoV-2 RNA concentrations using a 14-day sliding window, which yielded 301 values encompassing the entire follow-up period. These averages correlated strongly with 14-day cumulative notification rates reported by public health authorities (Pearson *r* = 0.962, *P* < 0.001). The estimated log-log regression slope was 1.011 ± 0.016, suggesting an approximately linear relationship between these two quantities over several orders of magnitude (Fig. 2A). A mean concentration higher than 1000 gc/L showed 95.6% sensitivity and 100% specificity as an indicator of a 14-day cumulative notification rate higher than 25 new cases per 100,000 inhabitants (Table 1). This disease incidence has been used in EU countries as a cutoff value below which restrictions to free population movements could be lifted.^27^ In turn, a mean concentration exceeding 10,000 gc/L showed 91.8% sensitivity and 95.1% specificity as an indicator of a 14-day cumulative notification rate above 350 new cases per 100,000 inhabitants, a high-concern incidence (Table 1). The correlation between 14-day cumulative notification rates and wastewater RNA concentrations was similarly high when medians were used for calculating the 14-day sliding window (r = 0.959, P < 0.001; Fig. S1A). We also found that the RT-qPCR results obtained with primers N1 and N2 were highly correlated (r = 0.952, P < 0.001) except for low-concentration samples, in which N2 tended to yield higher values than N1 (Fig. S1B). Similarly, there was a high correlation between data obtained from the two wastewater plant collectors (r = 0.951, P < 0.001; Fig. S1C).

**Figure 2.**
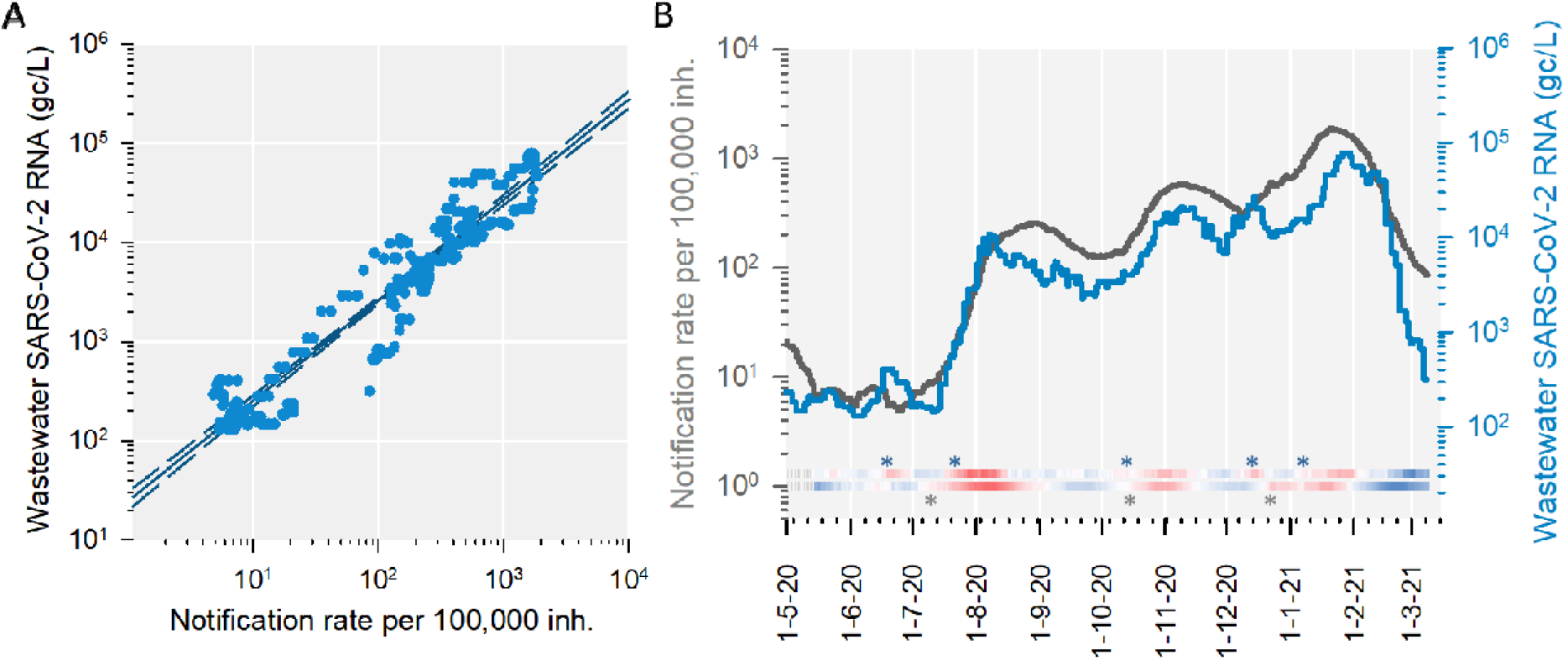
SARS-CoV-2 wastewater signal and notification rates over 14-day periods. Average log SARS-CoV-2 RNA concentrations in wastewater and cumulative case notification rates per 100,000 inhabitants are shown. These variables were calculated for each of 301 periods of 14 days using a one-day shifted sliding window. **A**. Least-squares log-log linear regression, showing 99% confidence intervals. Equation for the regression line: *y* = 1.011 *x* + 1.393. Goodness of fit: r^2^ = 0.925. **B**. Time series. The heat maps below the curves show the difference between consecutive 14-day periods (red: increasing, blue: decreasing) for wastewater data (top) and notification rates (bottom). Asterisks indicate the first day in which a significant increase between two consecutive 14-day periods was detected.

**Table 1.**
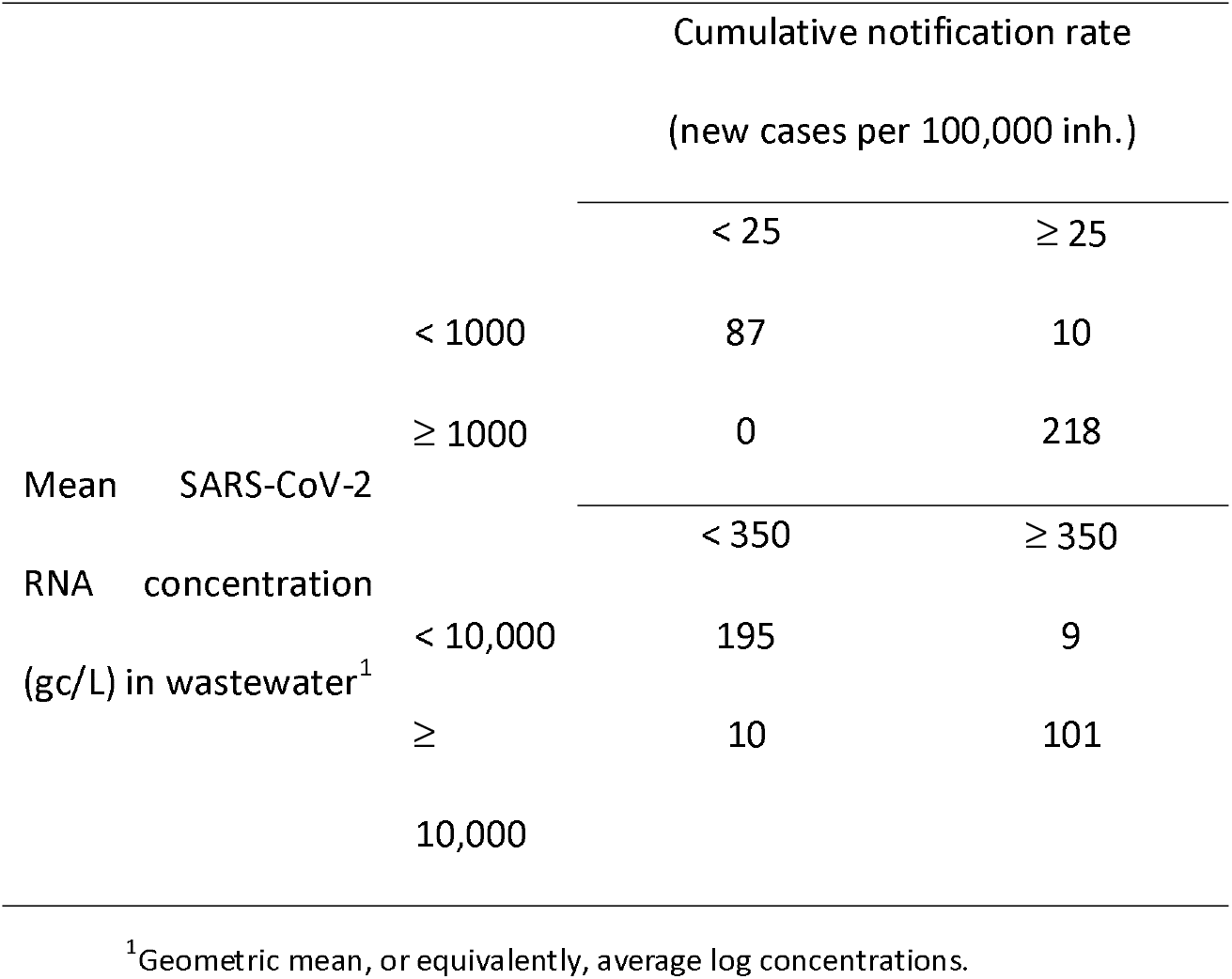
Categorical wastewater RNA concentrations versus disease incidences over 14-day periods.

|  |  | Cumulative notification rate<br>(new cases per 100,000 inh.) |  |
| --- | --- | --- | --- |
|  |  | < 25 | ≥ 25 |
| Mean SARS-CoV-2<br>RNA concentration<br>(gc/L) in wastewater<sup>1</sup> | < 1000 | 87 | 10 |
|  | ≥ 1000 | 0 | 218 |
|  |  | < 350 | ≥ 350 |
|  | < 10,000 | 195 | 9 |
|  | ≥ 10,000 | 10 | 101 |

<sup>1</sup>Geometric mean, or equivalently, average log concentrations.

To test the importance of taking composite samples, we also obtained individual (grab) samples from the same WWTP collectors on 91 different days, and performed 720 RT-qPCRs from these individual samples accordingly. The correlation between 14-day cumulative notification rates and average log SARS-CoV-2 RNA concentrations was much lower than for composite samples (r = 0.398, P < 0.001; Fig. S1D). This illustrates the importance of taking representative wastewater samples, although we note that the time period for this analysis was shorter (July 16, 2020 to March 9, 2021) and encompassed less variation in SARS-CoV-2 declared cases.

We also obtained composite samples from a single collector in a smaller WWTP (Quart-Benàger), which processes sewage from approximately 140,000 inhabitants, and performed 480 RT-qPCRs from samples taken on 120 different days (April 16, 2020 to March 9, 2021). We found a relatively high correlation between 14-day cumulative notification rates and average log SARS-CoV-2 RNA concentrations (r = 0.868, P < 0.001; Fig. S2A), although lower than for the larger Pinedo WWTP.

We then analyzed Pinedo WWTP data using a 7-day sliding window. Average log SARS-CoV-2 RNA concentrations correlated highly with 7-day cumulative notification rates, again exhibiting a linear relationship over several orders of magnitude (log-log Pearson r = 0.925, P < 0.001; slope = 1.009 ± 0.023; Fig. 3A). A mean concentration above 1000 gc/L showed 95.2% sensitivity and 95.7% specificity as an indicator of a 7-day cumulative notification rate higher than 12.5 new cases per 100,000 inhabitants, whereas a mean concentration exceeding 10,000 gc/L showed 88.6% sensitivity and 91.2% specificity as an indicator of more than 175 new cases per 100,000 inhabitants (Table 2). Therefore, wastewater analysis over weekly periods also provided a reliable tool, and could be used for early warning of SARS-CoV-2 spread.

**Figure 3.**
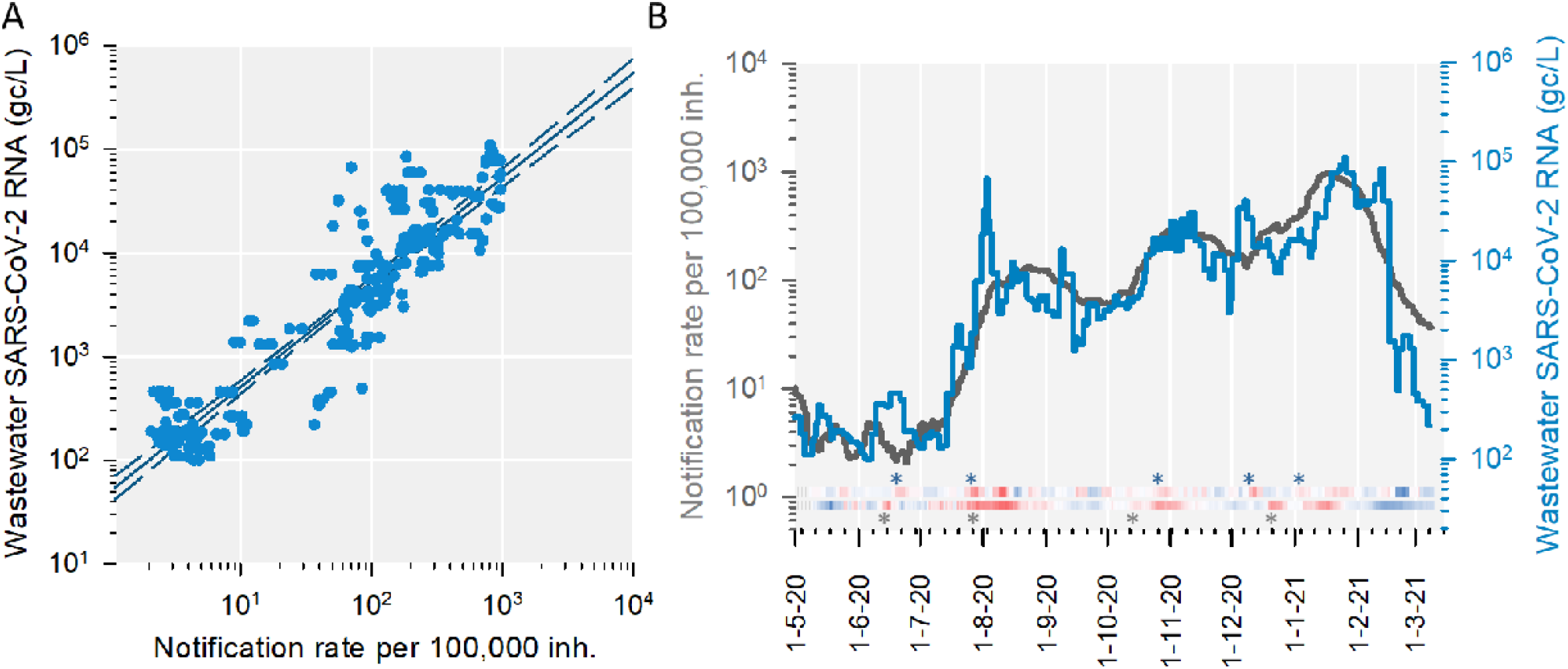
SARS-CoV-2 wastewater signal and notification rate over 7-day periods. Average log SARS-CoV-2 RNA concentrations in wastewater cumulative case notification rates per 100,000 inhabitants are shown. These variables were calculated for each of 322 periods of 7 days using a one-day shifted sliding windows. **A**. Least-squares log-log linear regression, showing 99% confidence intervals (*y* = 1.009 *x* + 1.699, r^2^ = 0.856). **B**. Time series. The heat maps below the curves show the difference between consecutive 7-day periods (red: increasing, blue: decreasing) for wastewater data (top) and notification rates (bottom). Asterisks indicate the first day in which a significant increase between two consecutive 7-day periods was detected.

**Table 2.** Categorical wastewater RNA concentrations versus disease incidences over 7-day periods.

|  |  | Cumulative notification rate<br>(new cases per 100,000 inh.) |  |
| --- | --- | --- | --- |
|  |  | < 12.5 | ≥ 12.5 |
| Mean SARS-CoV-2<br>RNA concentration<br>(gc/L) in wastewater<sup>1</sup> | < 1000 | 88 | 11 |
|  | ≥ 1000 | 4 | 219 |
|  |  | < 175 | ≥ 175 |
|  | < 10,000 | 198 | 11 |
|  | ≥ 10,000 | 19 | 93 |

<sup>1</sup>Geometric mean, or equivalently, average log concentrations.

We further investigated the ability of wastewater analysis to detect outbreaks by plotting side by side the time series of 14-day log SARS-CoV-2 RNA average concentrations and cumulative notification rates (Fig. 2B). Three waves of cases occurred in July, October and December 2020 in the region under study, all of which were clearly identified in wastewaters. By comparing consecutive 14-day periods, the first statistically significant increase in cases took place on July 9, 2020 (i.e. comparing June 12-25 versus June 26 - July 9; t-test using log transformed data: P = 0.022). In contrast, the first significant elevation of wastewater RNA concentrations was detected on June 18 (P = 0.013), followed by a decrease and, then, a more pronounced spike first detected on July 21 (P = 0.008). For the second wave, a significant increase in declared cases was first seen on October 14 (P = 0.042), whereas wastewater analysis first revealed such increase on October 13 (P = 0.032). The third wave of declared cases was first detected on December 22. Again, wastewater RNA concentrations increased before declared cases (first detected on December 13, P = 0.008) then decreased transiently, and rebounded more strongly in January, 2021. Finally, a decline in SARS-CoV-2 incidence and wastewater RNA concentrations started in February 2021 and continued until the end of the follow-up period.

Using a 7-day window, we also found good agreement between the time series of cumulative notification rates and average log wastewater RNA concentrations, although the later were noisier, as expected (Fig. 3B). With this approach, the first outbreak was detected on July 20 for declared cases (t-test: P = 0.027) and on July 19 for wastewaters (P < 0.001). Again, this was preceded by a transient peak in both wastewaters and declared cases between June 4 and 11 (P < 0.05). The second outbreak became significant for declared cases on October 8 (P = 0.046) and on October 21 for wastewaters (P = 0.017). Interestingly, the 7-day wastewater analysis detected another transient, marginally significant bout preceding this outbreak (September 10, P = 0.061). The third wave of declared cases was first detected on December 17 (P = 0.012). Again, we found that wastewater RNA concentrations increased before declared cases (December 6, P = 0.046), then decreased, and subsequently rebounded.

The transient peaks in wastewater SARS-CoV-2 RNA concentrations that preceded major outbreaks were confirmed by non-parametric statistics. By this approach, peaks starting on June 18 (Mann-Whitney U: P = 0.008) and December 13 (P = 0.003) were identified using a 14-day window. With a 7-day window, we obtained similar peaks (P = 0.012 on June 11 and P = 0.038 on December 6) as well as an additional marginally significant bout starting on September 10 (P = 0.057), consistent with the above results. To further test this transient-peak pattern, we analyzed data from the Quart-Benàger WWTP. Comparison of 14-day periods revealed significant bouts starting on June 13 (t-test: P = 0.007) and December 8 (P = 0.020) that preceding major outbreaks. We also found increased SARS-CoV-2 RNA concentrations during September 7-14, compared to August 23-31 (P = 0.034), thus supporting the presence of a short peak preceding the October wave.

## Discussion

Our analysis encompassing a period of major epidemic concern in the Spanish city of Valencia reveals that wastewaters can be used as a reliable indicator of COVID-19 disease incidence. We find that wastewater analysis is particularly useful for monitoring incidence values on the order of 25 new cases per 100,000 inhabitants over a 14-day period, or equivalently, 12.5 cases over a 7-day period. For these values, wastewater RNA concentrations achieve a specificity and a sensitivity greater than 95% using an RNA cutoff value of 1000 gc/L. We also estimated that the detection limit of the technique is on the order 100 gc/L, which should correspond to approximately two new cases per 100,000 inhabitants over a 7-day period.

To what extent these quantitative data can be extrapolated to other scenarios remains to be addressed. We have shown that compound sampling is essential for achieving a strong correlation between wastewater RNA concentration and declared cases, and that systematic sampling of large treatment plants should yield more accurate data. Additional factors that could influence the results include the sample concentration method, the RT-qPCR reagents used, and features of the local sewage system, among others. In addition, viral shedding through feces could vary depending on virological factors. For instance, certain SARS-CoV-2 variants might exhibit differences in organ tropism compared to the B lineage predominant in the region of Valencia over the course of this study, which was subsequently replaced by variant B1.1.7 during March 2021. Vaccination might also affect the association between wastewater RNA concentration and declared cases, since vaccination programs modified the age composition of the infected population and, potentially, could also alter viral shedding patterns.

Contrarily to previous suggestions,^17,20,23,28^ we did not find that wastewater analysis could systematically anticipate incidence peaks. Yet, intriguingly, transient elevations of wastewater viral RNA concentrations were detected approximately one month before major epidemic waves. Importantly, these peaks went unnoticed when looking at declared cases, or were very mild. We speculate that these cryptic peaks took place among subpopulations mainly constituted by asymptomatic carriers who failed to be diagnosed, particularly young individuals. These episodes should have been restricted to a specific time period or to certain communities that presumably became infected and then cleared the virus. For instance, this pattern might be due episodic population movements, such as travel or increases in social activity. Potentially, the transient peaks could have ignited viral transmission in the general population, causing major outbreaks in the weeks that followed.

Major lockdown measures were enforced in the region in March, 2020, allowing control of the first outbreak, which is not addressed in this study.^12^ On May 28, 2020, there was a major relaxation of certain anti-COVID-19 measures in the Valencian region, which included the opening of indoor spaces such as restaurants, sports centers and shopping malls, and had a measurable effect on cell phone-based population movement records (Fig. S3). This might have led to a reactivation of viral spread, mainly among younger people. Then, travelling between provinces or countries was allowed in June 21, matching the start of school holidays. This series of events could explain the observed evolution of SARS-CoV-2 RNA concentration in wastewaters. School classes were resumed on September 7, and viral RNA concentration in wastewaters increased precisely at that time, albeit weakly. It is thus possible that hidden viral spread during this period preceded the second major outbreak in October. Concerning the third transient peak observed in wastewaters prior to Christmas period, possible causes include the increase in commercial activity starting around November and the vacation period from December 5 to 8, during which indoor social activities could have increased, although these events did not appreciably impact population movement records (Fig. S3).

In conclusion, this study shows that SARS-CoV-2 population spread can be monitored reliably over a wide range of incidence values using wastewater analysis, and that wastewater viral RNA quantitation is a sensitive and specific indicator of early viral spread. Although we did not observe that this approach could systematically anticipate disease incidence peaks, we noticed that such peaks were preceded by transient elevations of RNA concentration in wastewaters. This finding deserves further examination and could potentially reveal novel aspects of COVID-19 epidemiology.

## Methods

### Wastewater sampling

Wastewaters from the Pinedo and Quart-Benàger WWTP in the metropolitan area of Valencia (Spain) were sampled (200 mL) by grabbing or taking 4 h composite samples between 7 and 11 am. Samples were kept at 4 °C and processed in the laboratory within 24 h of collection. Before processing, waters were spiked with 10^5^ plaque forming units of TGEV as an internal control. Then, samples were spun at 3000 × g for 10 min at 4°C, and the supernatant was filtered through 0.45 µm pore filters to remove large debris. Subsequently, 35 mL of filtered wastewater was centrifuged at 80,000 g, 3.5 h, 4°C to concentrate viral particles. Finally, the supernatant was removed and the pellet was resuspended in 500 µL phosphate-buffered saline (PBS).

### RNA extraction and SARS-CoV-2 RT-qPCR

RNA extraction was performed with the Nucleospin RNA Virus Kit (Macherey-Nagel) following the manufacturer instructions. RT-qPCR was carried out using the GoTaq^®^ Probe 1-Step RT-qPCR System (Promega) and the U.S. Center for Disease Control N1 and N2 primers set (2019-nCoV CDC EUA Kit, IDT). For each RT-qPCR run, calibration curves were performed using the 2019-nCoV_N_Positive Control provided in the kit. In addition, negative controls of the concentration, extraction and PCR process were included. Cycle threshold (Ct) values were used for calculating SARS-CoV-2 genomic copies per liter (gc/L) in the original sample based on calibration curves. Ct values < 40 were considered positive for SARS-CoV-2, as previously proposed.^29^ All PCRs were performed in duplicate. In-house primers were used for TGEV RT-qPCRs.

### Epidemiological data

Numbers of declared cases per day and per municipality were provided by the Conselleria de Sanidad Universal y Salud Pública (Generalitat Valenciana). The specific municipalities covered by each WWTP were provided by the Empresa Pública de Saneamiento de Aguas Residuales (EPSAR, Generalitat Valenciana). The analysis of local mobility in the city of Valencia was obtained from the publicly available population mobility data server of the Spanish Ministerio de Transporte, Mobilidad y Agenda Urbana.^30^

## Data Availability

The data that support the findings of this study are available from the corresponding authors, upon reasonable request.

## Acknowledgments

We thank Óscar Zurriaga (Conselleria de Sanitat Universal i Salut Pública, Generalitat Valenciana) for providing case notification rates, Augusto Montamarta and Ignacio García (Entitat de Sanejament d’Aigües, Generalitat Valenciana) for access to wastewater treatment plants, Ángel Jimenez, Pau Granell, Sonia Tristante and María José Tárrega for wastewater sampling, and Rocío Báguena for assistance with big data collection (Spanish Ministerio de Transporte, Mobilidad y Agenda Urbana). This work was funded by the Conselleria d’Agricultura, Desenvolupament Rural, Emergencia Climàtica i Transició Ecològica of the Generalitat Valenciana (project OTR2020-20593SUBDI to R.S.), the Instituto de Salud Carlos III (FONDO-COVID19 COV20/00210 to P.D-C.), CSIC (Salud Global CSIC 202020E292 to P.D-C), and the Spanish Ministerio de Ciencia e Innovación (Ramón y Cajal contract, Call 2019, to P.D-C.).

## Author contributions

Both authors contributed equally to the manuscript.

---

**Figure S1.**
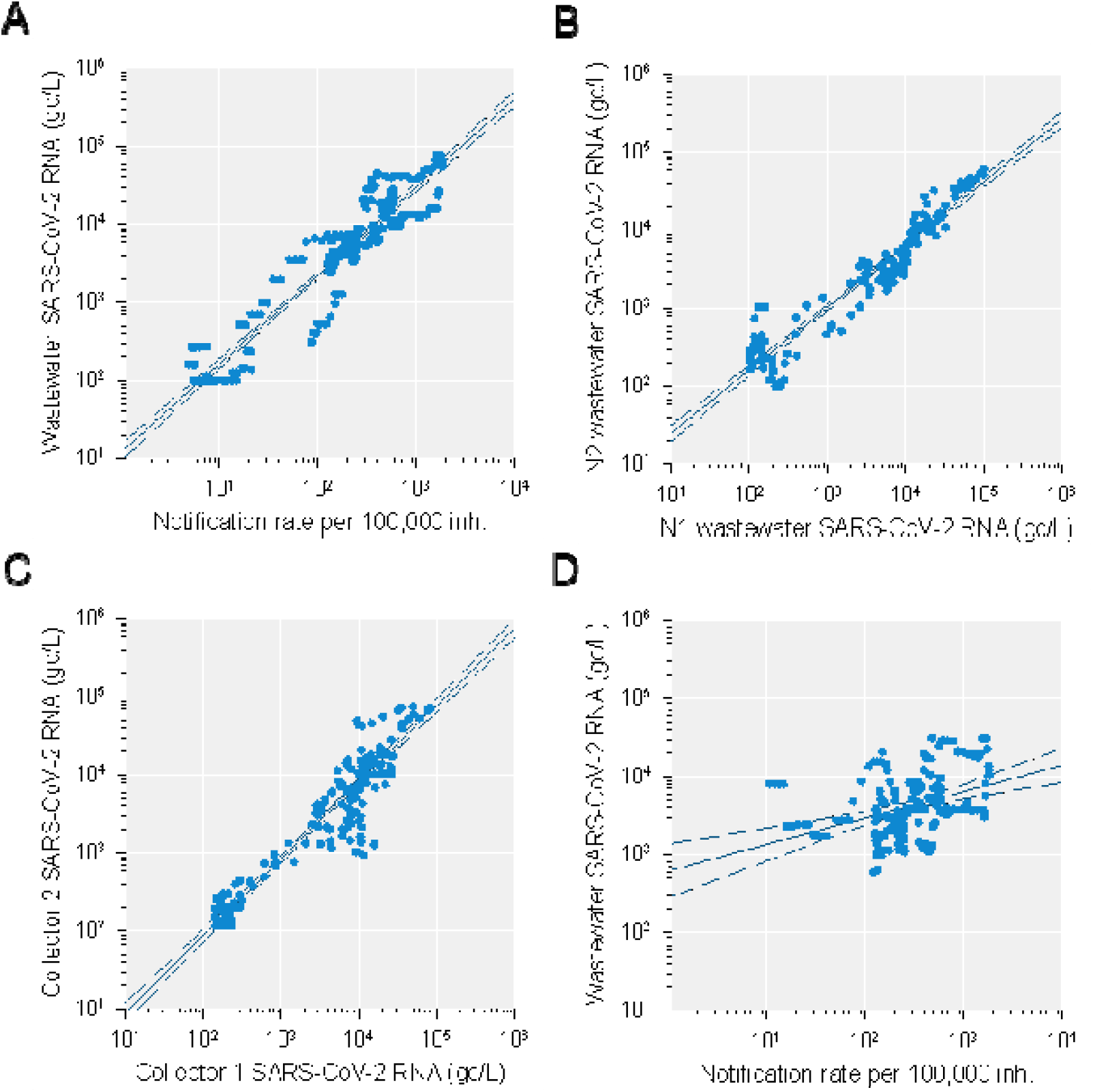
Influence of wastewater sampling strategy on SARS-CoV-2 quantitation. Viral RNA concentrations in wastewater and cumulative case notification rates per 100,000 inhabitants were calculated for 14-day periods using a one-day shifted sliding windows. Least-squares log-log linear regression lines and 99% confidence intervals are shown. **A**. Notification rate versus median viral RNA concentration in wastewaters. **B**. Average log viral RNA concentrations estimated using SARS-CoV-2 genome region N1 versus N2. **C**. Average log viral RNA concentrations estimated from the Pinedo WWTP collector 1 versus collector 2. **D**. Notification rate versus average log viral RNA concentration in wastewaters using individual instead of composite samples.

**Figure S2.**
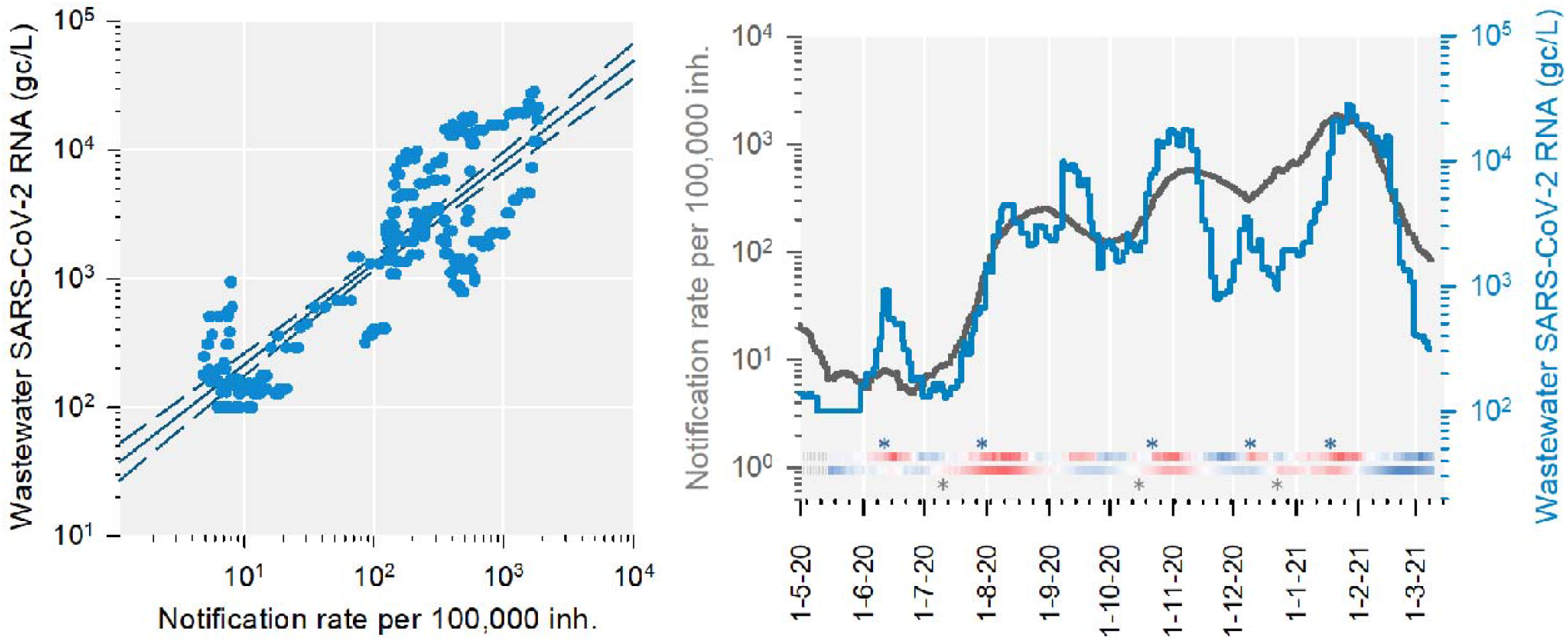
SARS-CoV-2 wastewater signal from a second treatment plant and notification rates over 14-day periods. Average log SARS-CoV-2 RNA concentrations in wastewater from Quart-Benàger WWTP and cumulative case notification rates per 100,000 inhabitants are shown. These variables were calculated for each of 301 periods of 14 days using a one-day shifted sliding windows. **A**. Least-squares log-log linear regression, showing 99% confidence intervals (*y* = 0.785 *x* + 1.547, r^2^ = 0.753). **B**. Time series. The heat maps below the curves show the difference between consecutive 14-day periods (red: increasing, blue: decreasing) for wastewater data (top) and notification rates (bottom). Asterisks indicate the first day in which a significant increase between two consecutive 14-day periods was detected.

**Figure S4.**
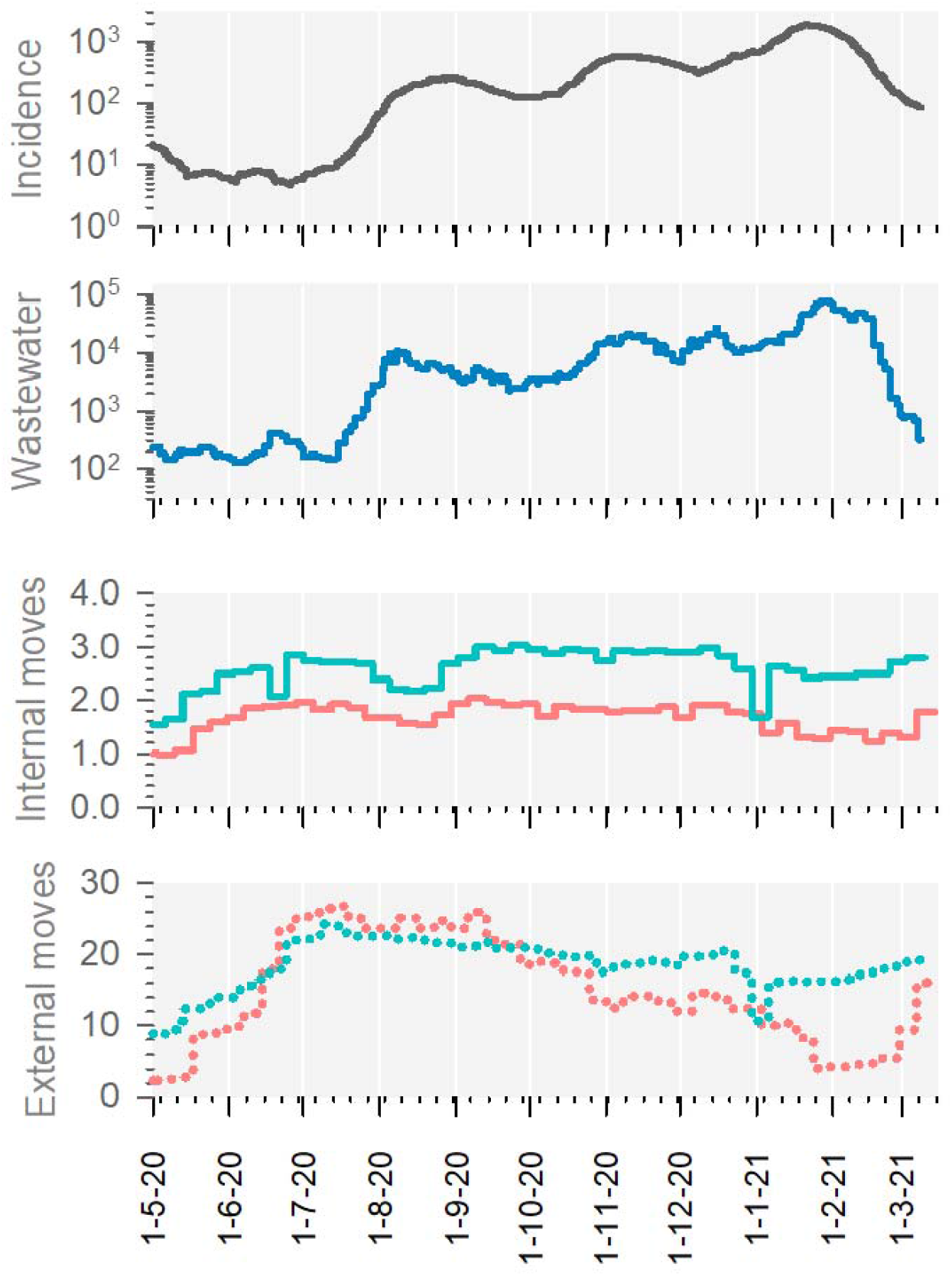
COVID-19 incidence, SARS-CoV-2 RNA concentration in wastewaters, and population mobility records. From top to bottom: 14-day cumulative declared cases per 100,000 inhabitants; average log SARS-CoV-2 RNA concentration (gc/L) over a 14-day sliding window; publicly available population movement records within the city of Valencia (Spain) on Wednesdays (green) and Sundays (red); population movements entering or departing from the city of Valencia (Spain) recorded on Wednesdays (green) and Sundays (red).

